# AI-Driven Surveillance of West Nile Virus in Maryland: Integrating Vector Identification with Environmental and Epidemiological Insights

**DOI:** 10.1101/2025.05.19.25327916

**Authors:** Khushi Anil Gupta, Hannah Markle, Alyssa Schultz, Brian Prendergast, Margaret Glancey, Autumn Goodwin, Tristan Ford, Roy Faiman

## Abstract

This study presents an integrated operational surveillance effort conducted in Anne Arundel County, Maryland, during the 2023 and 2024 mosquito seasons, revealing substantial variation in *Culex pipiens* s.l. abundance and West Nile virus (WNV) infection risk across time and space. In 2024, mosquito abundance and WNV-positive pools increased more than four- and five-fold, respectively, compared to 2023, underscoring the dynamic nature of local vector populations and arboviral transmission. Observationally, temperature was moderately predictive of mosquito abundance with a 1–2 week lag. Yet, correlational analyses only revealed a relationship between mosquito abundance and precipitation with a 3-week lag in 2023. Notably, ᐩthe temporal overlap between peak mosquito abundance and peak WNV infection was more synchronized in 2024, potentially heightening human transmission risk. These trends informed targeted vector control operations by the Maryland Department of Agriculture, demonstrating the importance of high-resolution, temporally responsive surveillance systems. This work also highlights the operational value of the IDX vector identification platform as part of a scalable entomological surveillance workflow. The mosquito imaging and identification capabilities of IDX supported sample triage, cold chain preservation, and molecular testing without delay. Its integration into the surveillance pipeline enhanced data integrity by enabling efficient specimen logging and species-level confirmation across thousands of samples. The combination of field collection, automated identification, and WNV testing illustrates a deployable model for responsive vector surveillance. This approach supports swift decision-making and demonstrates a modernized framework for mosquito control programs aiming to align operational capacity with climate-driven risk dynamics and advanced technologies.

## 1. Introduction

West Nile virus (WNV) remains a significant public health concern globally, including in the United States, where mosquitoes in the *Culex pipiens* Group (denoted here as *Cx. pipiens* s.l.) act as the principal vectors and wild birds act as reservoir hosts. Humans serve as dead-end hosts when these mosquitoes, having fed on infected birds, bite humans and transmit the virus, establishing an infection (Kilpatrick et al. 2005, Kilpatrick and Pape 2013). Mosquito population dynamics and vector competence are influenced by meteorological variables such as temperature and humidity (Shocket et al. 2020) and rainfall patterns affect aquatic habitats necessary for larval development, influencing adult mosquito densities (Gardner et al. 2012, Karki et al. 2016). In temperate climates for example, the timing of mosquito egg-laying and hatching is influenced by spring warming rates, linking climate variability to mosquito reproduction and subsequent vector dynamics (Burkett-Cadena et al. 2012, Shave et al. 2019). Typically, WNV transmission peaks in late summer, reflecting seasonal variations in mosquito abundance and virus amplification (Shocket et al. 2020).These interconnected environmental factors collectively shape mosquito populations, virus amplification in mosquito and avian hosts, and ultimately, the density of infected mosquitoes that determine WNV transmission risk to humans (Paz 2019, Reisen 2013). Therefore, understanding and mitigating WNV transmission requires robust surveillance systems that account for the complex interplay of such environmental factors. Likewise, extreme events such as heatwaves, hurricanes, droughts, and anthropogenic interference can all impact mosquito abundance, vector competence, and avian host populations (Shocket et al. 2020, Gardner et al. 2012, Karki et al. 2016, Albright et al. 2010, Paz 2019, Hermanns et al. 2023).

Mosquito pool testing is currently the most efficient and commonly used method for monitoring WNV in mosquito populations. This approach is used uniformly across health agencies and provides standardized infection rate data, making it a cornerstone of mosquito surveillance programs. This method relies on two essential indices, the minimum infection rate (MIR) and the vector index (VI), to provide complementary insights into infection dynamics. The MIR serves as a straightforward and critical early warning system for identifying areas with active WNV transmission. Conversely, the VI incorporates both mosquito abundance and infection prevalence to provide a comprehensive understanding of infection intensity and human risk, enabling public health and mosquito control agencies to prioritize resources by designing targeted interventions (Kilpatrick et al. 2005).

Mosquito control organizations (MCOs), agencies that are typically public-based and responsible for monitoring and mitigating nuisance mosquitoes, vector populations, and associated diseases, often rely on labor-intensive surveillance methods that struggle to provide timely and actionable insights (NACCHO 2017, 2024). An additional challenge faced by MCOs is a shortage of expertise in accurate mosquito identification. With medical entomologists in short supply (Harrington and Mader 2023), much of the work involves seasonal technicians who often have limited skills and experience in mosquito identification. This knowledge gap can result in misidentifications or delays in obtaining critical data, compromising the implementation of vector control. As accurate species identification is crucial to understanding vector competence and executing targeted interventions, this shortfall creates a significant bottleneck in public health efforts to manage WNV transmission. Moreover, shifting species distributions and the emergence of invasive vectors increasingly demand precise taxonomic identification to detect and respond to novel threats before they drive local disease transmission (Crowl et al. 2008, Early et al. 2016).

The emergence of automated vector identification tools offers a transformative solution to these challenges. The IDX tool (Vectech Inc., Baltimore, MD), an imaging device that provides a transformative artificial intelligence (AI)-based solution by harnessing computer vision and deep learning algorithms, assists technicians and public health professionals in streamlining mosquito identification and data curation. IDX is capable of analyzing specimens with efficiency and consistency similar to or superior to that of human technicians or expert entomologists (Brey et al. 2022, Gupta et al. 2024). IDX facilitates the monitoring of WNV, other vector-borne pathogen risks, and invasive or uncommon species by providing accessible mosquito identification capability in minimal time (Faiman et al. 2024). The bridge vector paradigm, which emphasizes the role of mosquitoes in bridging the virus between avian and human hosts, highlights the importance of understanding mosquito abundance and infection dynamics to predict human WNV infections (Kilpatrick et al. 2005). By providing insights into bridge vector diversity and abundance, this technology enables agencies to prioritize interventions and allocate resources effectively when coupling with infection rate data. This study showcases the utility of the IDX identification platform in supporting mosquito abatement efforts in central Maryland. Using data from Anne Arundel County collected during the summers of 2023 and 2024 for the Maryland Department of Agriculture (MDA), we illustrate how IDX supports rapid species identification and verification, enables robust data collection and reporting, maintains sample integrity for arboviral testing, and informs targeted mosquito control campaigns. By integrating IDX with weekly surveillance and environmental data, we highlight our findings of mosquito burden and WNV transmission in Anne Arundel County, Maryland.

## 2. Methods

### Surveillance Framework

Weekly collections were completed at 12 sites in Anne Arundel County, Maryland during the summers of 2023 and 2024 (Figs. S1-S2). Collections ran each year during the last full week of June (EpiWeek 26) through the last week of September (EpiWeek 39). Sites were selected by the Maryland Department of Agriculture, targeting areas proximal to water facilities where mosquito breeding may flourish or residential areas where vector-to-human interactions may occur. In 2024, two collection sites were replaced to increase representation across the county. CDC Gravid Traps (John W. Hock Company, Gainesville, FL) were utilized to standardize sampling and target the key WNV vector species *Cx. pipiens* s.l. Hay infusions were prepared by mixing 3 gal of warm water, 100 g of cut grass, and a 1/4 tsp of dry instant yeast (Lesaffre Corp., Milwaukee, Wisconsin) to a bucket. Separately, 1 tbsp of dry chicken manure pellets (Manures.com, West Covina, CA) were added to 1 gal of warm water. Both mixtures were allowed to incubate for four days prior to weekly collections. On the morning of collections, 2 cups of hay infusion were added to the chicken manure solution in gallon jugs. In the field, oviposition trap tubs were filled with 1 gal of hay infusion mixture to replicate stagnant water conditions preferred by ovipositing, gravid *Cx. pipiens* s.l. females (Lampman and Novak 1996). Traps were set at each collection site in the morning powered by a 6V lead-acid battery (John W. Hock Company, Gainesville, Florida) and collected after 24 hours. Trap collection bags were placed in a cooler lined with ice packs to maintain a cold chain necessary for minimizing RNA virus degradation and transferred to a -20°C freezer when collections were complete.

Specimens were sorted, identified to species and counted shortly after, while maintaining cold-chain conditions.

### IDX Workflow

All captured mosquitoes were identified and sorted by Vectech’s entomologists on each collection date. *Culex pipiens* s.l. females were processed immediately and pooled by collection site with up to 40 mosquitoes per pool. IDX imaging of all remaining specimens including damaged, rare, or unknown specimens to newly trained entomologists followed in order to log data and verify species and sex identifications (Figs. S3-S7) using the deployed algorithm described below. All IDX imaged and identified relevant female mosquito specimens were similarly pooled. Pools were submitted to the Maryland Department of Health Laboratories Administration, Division of Molecular Biology - Molecular Diagnostics/Bioterrorism (Baltimore, MD) on dry ice for RT-qPCR arbovirus screening which included assays for WNV, St. Louis encephalitis virus (SLEV), and Eastern equine encephalitis virus (EEEV). Cold chain integrity was maintained throughout the process.

### Vector Identification Model

IDX is a smart digital microscope designed for batch imaging and identification of mosquitoes and ticks (Brey et al. 2022, Achee, 2022) (Fig. S6). It is a cloud-connected lab device that streamlines vector surveillance data acquisition and curation, allowing a minimally-trained user to generate high quality, standardized species-level data on captured specimens (Fig. S6). IDX functions with a mature, deep convolutional neural network (CNN) trained on a large dataset of mosquito images to classify imaged mosquitoes based on visually distinguishable features (Goodwin et al. 2021, Brey et al. 2022). Algorithm versions v2.0.0, v2.0.1, v4.0.1, and v4.0.2 were used for these surveillance projects.

### Weather Data Integration

Daily temperature and precipitation data were extracted from Anne Arundel County weather station archives (ncei.noaa.gov 2023, 2024) to assess environmental factors driving mosquito population dynamics. Only one weather station in the county reported temperature data. Precipitation data was averaged from four weather stations representative of the study area and in closest proximity (miles) to collection sites for each year (Figs. S1-S2). Weather data from EpiWeeks 22-25, the four weeks preceding the collection period, were included to assess patterns of mosquito abundance correlated to previous weeks’ weather. Weekly average temperatures using minimum and maximum temperatures (°C) were calculated. Weekly cumulative precipitation (in mm) was averaged and error bars were calculated using the standard error of the mean to represent overall rainfall patterns across the study area.

### Data Scope and Analysis

Species diversity, abundance, and WNV prevalence were analyzed to guide MDA’s control campaigns in Anne Arundel County. Species diversity and abundance were represented by calculating the total captured mosquitoes per species across all traps collected over EpiWeeks 26-39 in 2023 and 2024. The abundance was calculated using the geometric William’s mean of collection numbers across all traps, applying log transformation to reduce variation, normalize distributions, and minimize the influence of extreme values driven by environmental, technical, or site-specific conditions. (Haddow 1959).

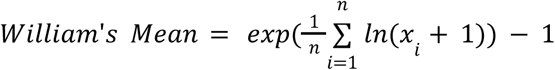

The minimum infection rate (MIR) of *Cx. pipiens* s.l. was calculated by dividing the total number of positive pools (Σ *positive pools*) by the total number of mosquitoes screened (Σ *mosquitoes screened*) based on standard WNV surveillance methods (CDC.gov 2025). The MIR is used to estimate infection rates by assuming that each positive pool contains only one infected individual, providing a reliable lower bound of the true infection rate, especially when infection rates are low or pool sizes are small (Gu et al. 2008).

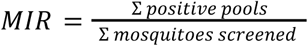

The vector index (VI) of *Cx. pipiens* s.l. was calculated by multiplying the average number of mosquitoes per trap night (µ_*mosquitoes per trap night*_) by the estimated proportion of mosquitoes infected (*MIR*) (CDC.gov 2025). The VI estimates the average number of WNV-infected mosquitoes per trap night by combining mosquito abundance with infection prevalence, providing a measure of potential transmission risk (Gujral et al. 2007).

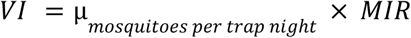

We conducted statistical and graphical analyses for this study using Tableau and Python.

To further assess the relationship between both cumulative precipitation and average temperature with William’s mean mosquito abundance, we performed a correlation study by calculating the Pearson correlation coefficient and p-value for both 2023 and 2024. To account for time-lagged effects, abundance lag ranging from 0-to-5 weeks was assessed based on the mosquito life cycle and previous findings correlating mosquito abundance to environmental factors over these periods of time (Buckner et al. 2011, Gardner et al. 2012, Karki et al. 2016) (Table S1).

## 3. Results

### Species Diversity and Abundance

A total of 4,523 female mosquitoes were collected and identified over the course of the season in 2023 and 17,532 in 2024. Of these, *Cx. pipiens* s.l. represented 3,882 (86%) and 16,244 (93%) in 2023 and 2024, respectively (Figs. 1-2, left). In 2023, ten other species were collected including *Aedes albopictus* (n=309)*, Ae. j. japonicus* (n=178)*, Anopheles quadrimaculatus* (n=86)*, Ae. triseriatus* (n=31)*, Cx. erraticus* (n=20)*, An. punctipennis (n=11), Ae. vexans* (n=2)*, Orthopodomyia signifera* (n=2), *An. crucians* s.l. (n=1), and *Cx. salinarius* (n=1) (Fig. 1). In 2024, ten other species were collected including *Ae. albopictus* (n=718)*, Ae. j. japonicus* (n=336)*, Ae. triseriatus* (n=74)*, An. quadrimaculatus* (n=67), *Cx. erraticus* (n=46), *Cx. restuans* (n=25), *An. punctipennis* (n=17), *Ae. vexans* (n=2)*, Psorophora ferox* (n=2), and *An. barberi* (n=1) (Fig. 2). The greatest abundance of non-*Culex* mosquitoes was *Ae. albopictus* in both years (48% and 55% of total other species in 2023 and 2024, respectively) followed by *Ae. j. japonicus* (approximately 27% in both years) (Figs. 1-2).

**Figure 1.**
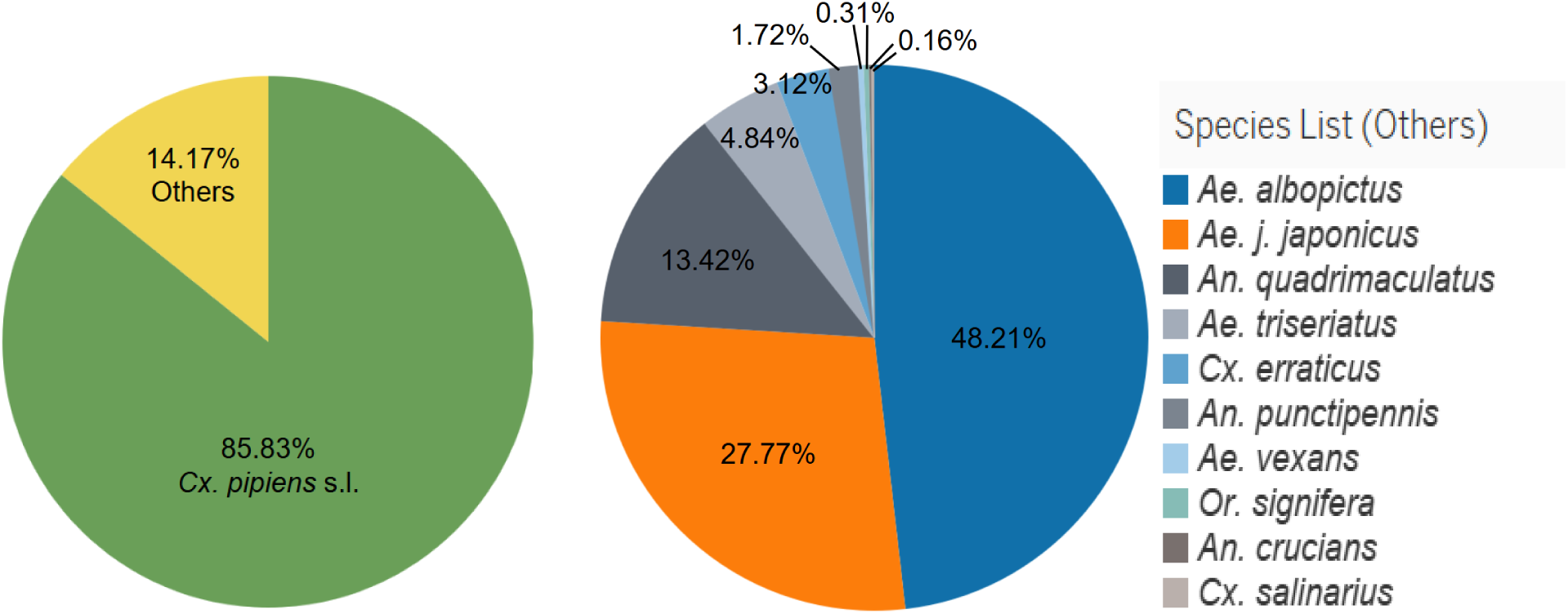
Species diversity and abundance (2023). *Culex pipiens* s.l. abundance (green, left panel) versus the ten other species combined (yellow, left panel) and their expanded breakdown (right panel).

**Figure 2.**
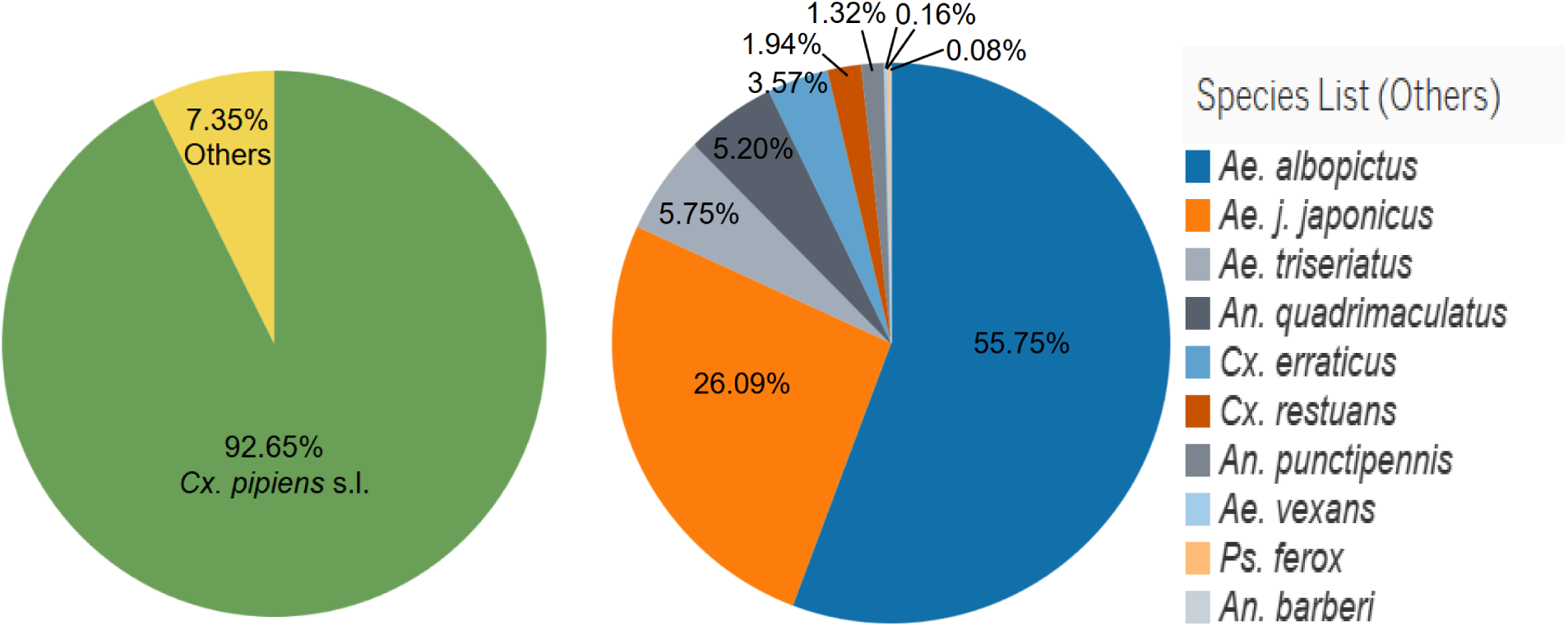
Species diversity and abundance (2024).*Culex pipiens* s.l. abundance (green, left panel) versus the ten other species combined (yellow, left panel) and their expanded breakdown (right panel).

### Environmental Drivers of Mosquito Dynamics

Weather conditions, particularly temperature and precipitation, were incorporated into our analysis to identify mosquito population trends and environmental factors driving vector proliferation. In 2023, temperatures gradually rose throughout the season, peaked during EpiWeek 36, and dropped off at the tail end of the season (Fig. 3). Conversely, temperatures peaked early in the season in 2024, remained high throughout the season, and gradually decreased across the season (Fig. 4). Rainfall followed somewhat similar patterns between years, with little precipitation in the weeks leading up to the surveillance period, peak rainfall during mid-season, a dry mid-to-late summer, and rainfall picking back up in late-September (Figs. 3-4). Two weeks early in the season during 2023 (EpiWeeks 25-26) showcased substantial cumulative precipitation (Fig. 3). In relation to mosquito abundance, warmer weather and/or increased precipitation occasionally coincided with subsequent spikes in *Cx. pipiens* s.l. abundance based on preliminary observations (Figs. 3-4). Abundance peaked during EpiWeek 28 in 2023 (Fig. 3) and EpiWeek 27 in 2024 (Fig. 4). Less extreme peaks were observed at EpiWeeks 30 and 35 in 2023 (Fig. 3), and EpiWeeks 35 and 36 in 2024 (Fig. 4). The lowest abundance was observed in EpiWeek 39 in 2023 (Fig. 3) and EpiWeek 26 in 2024 (Fig. 4), preceded by some of the lowest temperatures and cumulative rainfall of both years. Notably, results for EpiWeek 26 in 2024 may have been skewed due to multiple trap malfunctions, allowing for only 5 successfully functioning traps out of 12. Further, we examined correlations between weather and mosquito abundance in both years. Positive correlations were found between abundance and precipitation with a 3-week lag in 2023 (r = 0.564, p = 0.045), and temperature with a 0-week lag in 2023 (r = 0.577, p = 0.389), although the latter was not statistically significant (Table S1). A moderate positive correlation (r = 0.474) was seen between abundance and temperature with a 1-week lag in 2023, although not statistically significant (p = 0.102) (Table S1). Moderate correlations between abundance and precipitation with 3- and 5-week lags were seen in 2024 (r = 0.302 and r = 0.349, respectively), although they were not statistically significant (p = 0.294 and p = 0.242). Positive or significant correlations were not found in the remaining analyses (Table S1).

**Figure 3.**
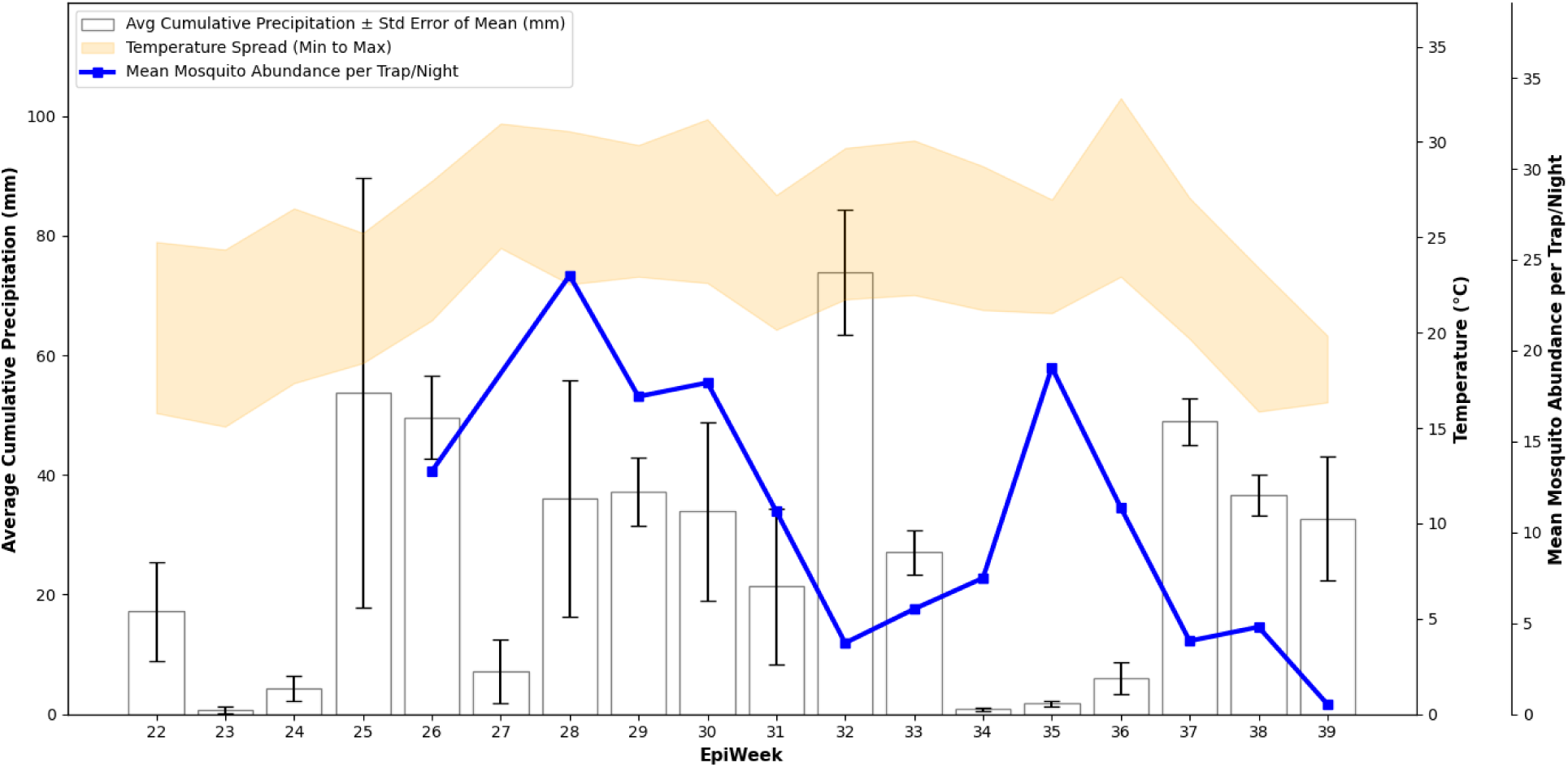
Mosquito abundance 2023. Adult female *Cx. pipiens* s.l. (geometric mean, blue line) collections for EpiWeeks 26-29 (June 30, 2023 - September 24, 2023) with average cumulative precipitation (mm, ± standard error, white bar graphs) and temperature (°C) spread (minimum to maximum, yellow shaded area) for EpiWeeks 22-39 (May 28, 2023 - September 30, 2023).

**Figure 4.**
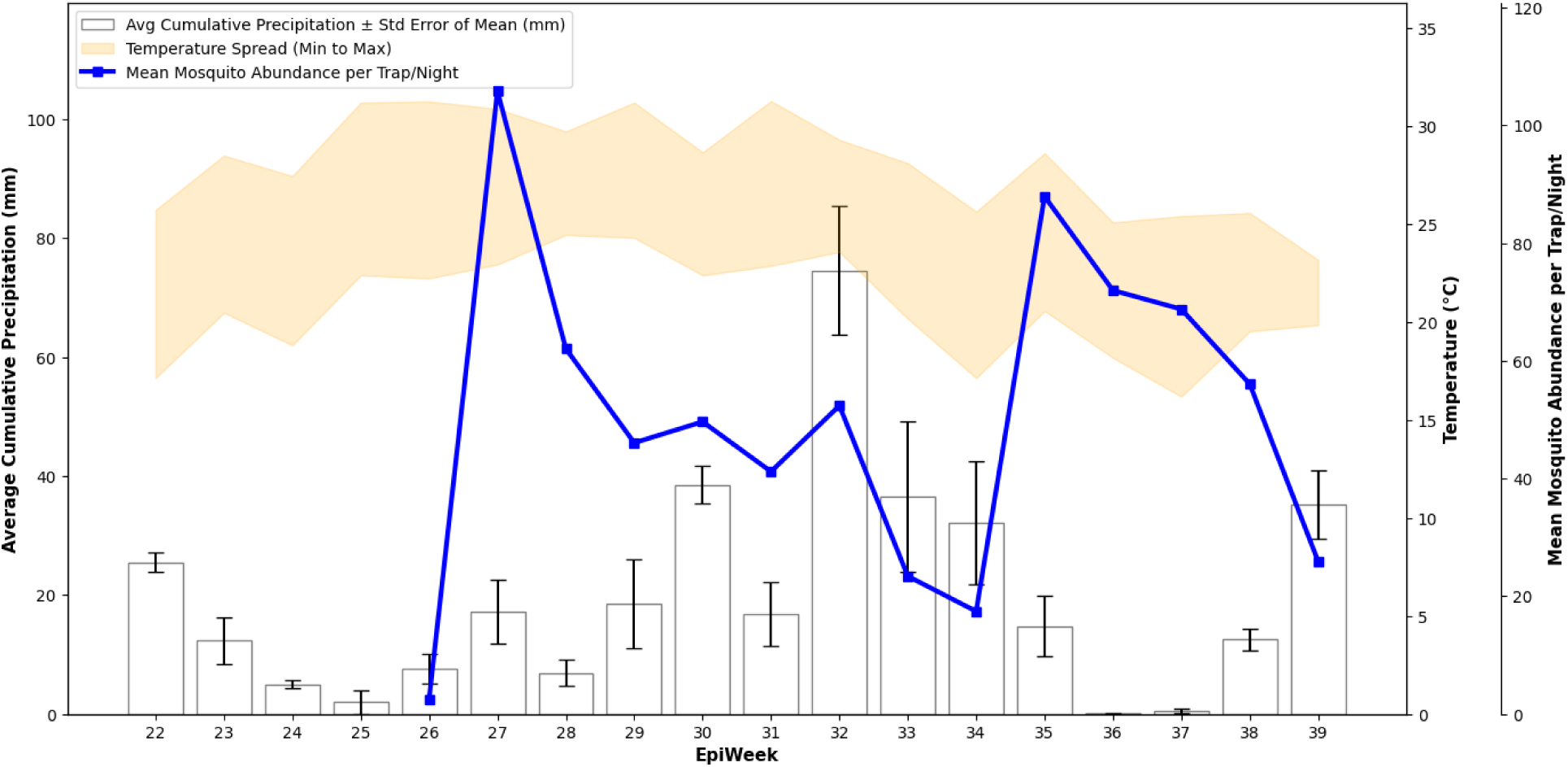
Mosquito abundance 2024. Adult female *Cx. pipiens* s.l. (geometric mean, blue line) collections for EpiWeeks 26-29 (June 26, 2024 - September 25, 2024) with average cumulative precipitation (mm, ± standard error, white bar graphs) and temperature (°C) spread (minimum to maximum, yellow shaded area) for EpiWeeks 22-39 (May 26, 2024 - September 28, 2024).

### West Nile Virus Mosquito Infection Rate

Molecular arboviral screening revealed trends in WNV infected pools over the mosquito seasons. A total of 211 pools were tested for pathogens in 2023 and 824 pools in 2024. WNV was detected in 35 pools of *Cx. pipiens* s.l. in 2023 and 183 pools in 2024. WNV was detected in 1 pool of *An. punctipennis* in 2024. No SLEV or EEEV was detected. The VI peaked during EpiWeek 29 in 2023 and EpiWeek 30-31 in 2024 (Fig. 5). Minimum infection rates followed suit in 2024, with the peak MIR occurring in EpiWeek 30-31. The peak MIR occurred later in 2023 during EpiWeek 33 (Fig. 5).

**Figure 5.**
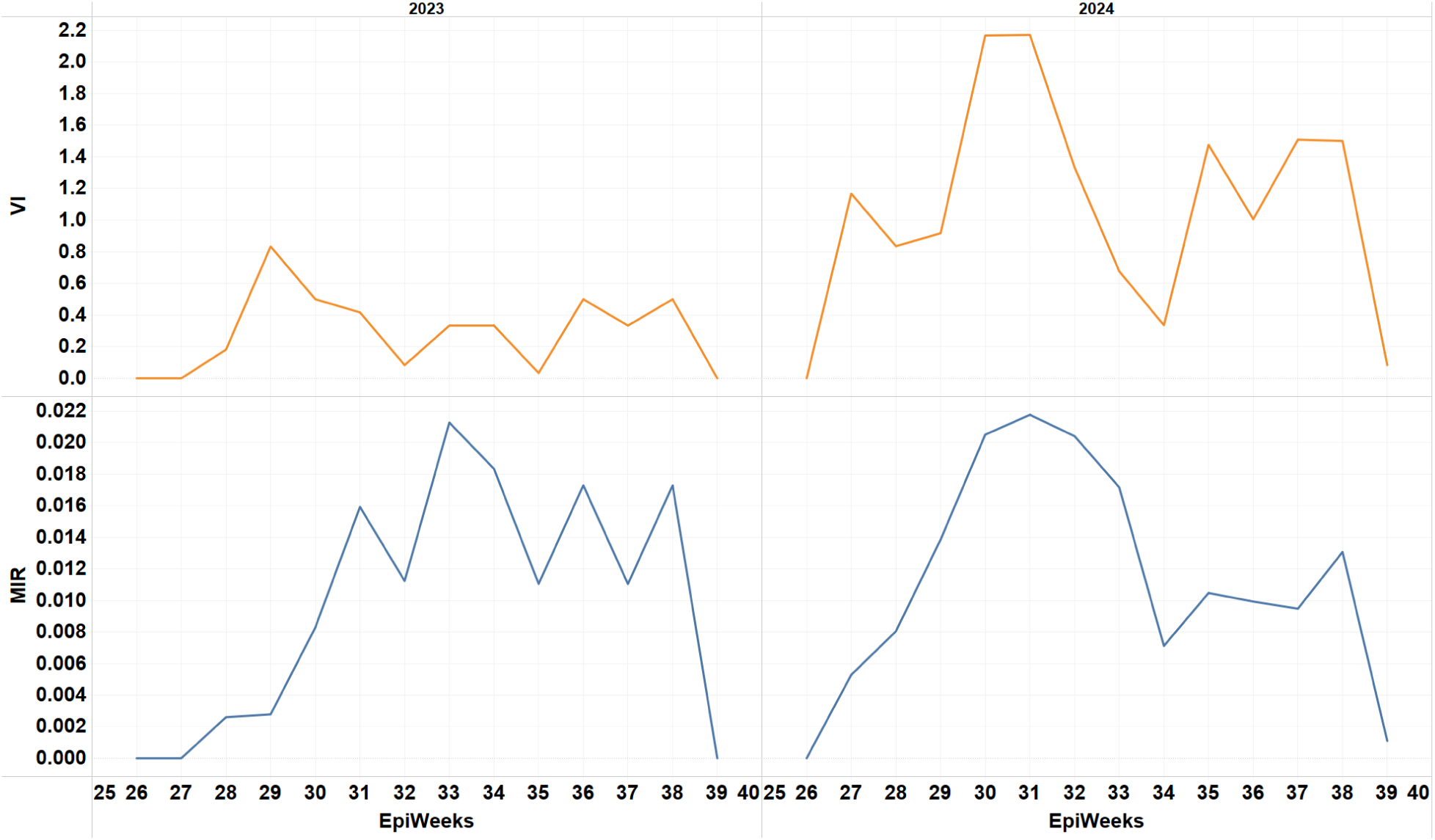
WNV surveillance indices for 2023-2024. Graphs depict trends in vector index (VI) (orange line, top left and right) and minimum infection rate (MIR) (blue line, bottom left and right) during the 2023 (top left and bottom left) and 2024 (top right and bottom right) surveillance seasons (EpiWeeks 26-39).

## 4. Discussion

### IDX as a Comprehensive Surveillance Tool

Integrating IDX into the West Nile virus (WNV) mosquito surveillance program proved valuable for supporting vector identification and streamlining data tracking. Traditional methods, such as inspecting individual features or using dichotomous keys, are time-consuming and prone to inaccuracies, with variability between labs and individuals (Goodwin et al. 2021, Jourdain et al. 2018). IDX helped overcome these challenges by providing rapid species identification for batches of specimens, allowing entomologists to maintain sample quality and reduce the time between collection and arboviral testing—an important metric for WNV forecasting (DeFelice et al. 2019). Additionally, IDX served as a training tool for interns, helping them close gaps in taxonomic identification skills by offering high-quality specimen images for reference. It also supported seasonal technicians and early-career entomologists in refining their identification abilities, particularly when classifying damaged or rare specimens, and in flagging unknown species. A study by Placer County Mosquito and Vector Control District confirmed that IDX outperformed seasonal technicians with limited experience (7-22 months) in both speed and accuracy (Hubble et al. pending publication). In this setting, IDX primarily functioned as a confirmatory identification tool and a digital logging platform, which facilitated data collection, reporting, and reduced labor costs.

Further, IDX is a scalable tool with a workflow applicable to many arboviral and vector surveillance efforts. Although WNV was the sole arbovirus detected in our mosquito pools out of those screened for, the vector identification capabilities of IDX prove to be a useful tool in timely surveillance of any pathogen. In light of recent locally acquired malaria cases in Maryland, the emerging Oropouche virus, and sporadic Dengue virus outbreaks in the US, quick and efficient vector identification capabilities for MCOs and abatement districts remain crucial (Duwell et al. 2023, Guagliardo et al. 2024, Wheaton et al. 2025). With the threat of disease outbreaks and the potential for invasive vectors to encroach on the region, early detection, made possible here through IDX, is essential to spur rapid vector control efforts.

Subsequent to exploring mosquito diversity and abundance, effective reporting and communication of the data become essential. Public health agencies and MCOs must develop integrated systems for sharing of vector surveillance, weather, and health data between agencies. Currently, reports generated on the IDX dashboard can be leveraged to visualize and share geographical, chronological, and taxonomical vector information. By incorporating IDX reports into health reporting or databases, agencies may gain a “bigger picture” view of WNV surveillance, allowing for enhanced and refined mitigation efforts.

### Integration of Environmental Data

Through weekly surveillance efforts during the summers of 2023 and 2024 in Anne Arundel County, we were able to track *Cx. pipiens* s.l. abundance at 12 strategic locations, gaining critical insights into the population dynamics of one of the primary vectors for WNV. Continuous data collection enabled identification of seasonal mosquito abundance peaks and facilitated the delineation of hotspots and immediate vector control interventions in surrounding areas. For instance, we noticed upward trends in trap abundance in the weeks following higher temperatures, likely due to accelerated breeding cycles and shorter larval development periods. However, strong correlations were not found between the two. Similarly, mosquito abundance appeared to trend upward in the weeks following greater cumulative precipitation. We hypothesize this was due to rainfall patterns influencing the availability of breeding habitats such as standing water. However, coinciding with or immediately post-peak cumulative precipitation, drops in mosquito abundance were seen compared to weeks prior. This could be due to major rain events washing out existing breeding sites as seen in other studies (Ruiz et al. 2010, Koenraadt and Harrington 2008, Geery and Holub 1989). Regardless, with many collection sites situated in close proximity to water treatment plants, breeding site availability may not be limited to major precipitation events alone. Supporting this notion, our correlation study largely did not reveal a strong relationship between environmental factors and mean mosquito abundance (with the exception of a positive correlation with a 3-week lag in 2023), suggesting that mosquitoes may not immediately respond to microclimatic events. Instead, more constant and larger breeding grounds might play a greater role, as well as our analysis utilizing the geometric mean across all traps which could dilute the relationship per site to the weather conditions. Further, sporadic instances of trap components being knocked over due to outside factors like animals, people, or weather events compromised trap integrity and consequently, mosquito catch numbers in both 2023 and 2024. While our data represents a comprehensive analysis of Anne Arundel County vector abundance, such limitations should be considered when interpreting results. Still, in forthcoming mosquito seasons, further analyzing weekly weather patterns may help track and predict mosquito surges in the following weeks, guiding preemptive control measures.

Refining our assessment of the vector species composition in the region will help us understand disease dynamics over time. Some mosquitoes, their associated pathogens, and vectorial capacity are expected to proliferate with temperature increase, while others are predicted to dwindle (Shocket et al. 2020, Brüssow and Figuerola 2025). This variability, coupled with Anne Arundel County’s situation within a mid-Atlantic temperate region warrants the continued evaluation of the intersection of weather and WNV cases over time. Future projects in Anne Arundel County could incorporate climate-based models similar to the study by Gong, DeGaetano, and Harrington (2010) to predict abundance of northeastern US vectors and associated disease burden. Overall, by continuing to closely track *Culex* vector abundance and compiling past and present surveillance results, we plan to explore longitudinal data to model long-term impacts of climate change on mosquito dynamics and WNV risks.

### Public Health Implications

Our data illustrating mosquito abundance, MIR, and VI from EpiWeeks 26 to 39 (July to September) of 2023-2024 offers insights for public health decision-making. In 2023, mosquito abundance remained low and stable with a modest peak around EpiWeek 28, while MIR gradually increased and peaked near EpiWeek 33. The weak correlation between the two (r = 0.37, p = 0.16) suggests infection dynamics were influenced by other factors such as pathogen introduction timing, mosquito age structure, or environmental conditions (McMillan et al. 2023). By contrast, 2024 saw a sharper, earlier abundance peak around EpiWeek 27, with MIR closely tracking that pattern (r = 0.49, p = 0.053), indicating a more synchronized transmission cycle.

This pattern aligns with McMillan et al. (2019), who found that WNV amplification was strongest when the dominant vector, *Cx. quinquefasciatus*, overlapped with the avian breeding season under favorable environmental conditions. Further supporting this, Humphreys et al. (2021) showed that WNV risk is shaped by complex interactions among climate, host communities, and demographic factors, emphasizing that vector abundance alone is an incomplete predictor of transmission. Together, these findings highlight the importance of integrating abundance and infection data with ecological and demographic context moving forward to guide timely, targeted public health interventions.

Despite expecting similar trends between 2023 and 2024 due to similar climatic conditions and localities, a nearly 4-fold increase in abundance and arboviral pool submissions and a 5-fold increase in positive WNV pools were observed in 2024 compared to the prior year. Species diversity remained largely consistent across both years. However, due to the limited scope of our surveillance (one trap night per week at only 12 sites in a 588 square mile county), these findings cannot be generalized to the broader region. Variations in abundance may be influenced by a number of factors such as hardware function, differences in avian populations, breeding site availability, greater experience by technicians, or more refined and improved surveillance protocols, though these remain speculative. Overall, mosquito counts alone may not sufficiently predict infection risk, emphasizing the importance of integrating both abundance and pathogen data into surveillance schemes.

Importantly, surveillance data guided MDA in identifying hotspots and initiating targeted control efforts. In response to WNV detection, marked by at least two positive mosquito pools, MDA implemented ultra-low volume (ULV) permethrin spraying within 1.2 km of collection sites in both years, with additional larvicide applications (granules or briquets) deployed in 2024.

However, the impact of these abatement efforts was not immediately reflected in subsequent surveillance data. This could be attributed to several factors, including intervention timing, environmental conditions, and critically, the spatial and temporal disconnect between mosquito collection sites and areas of operational control. As shown by McMillan et al. (2023), mosquito populations within 10-20 km of each other exhibited synchronized fluctuations in abundance, while arbovirus detection rarely synchronized beyond five km. This discrepancy highlights how arbovirus transmission tends to operate at more localized scales, potentially limiting the ability of trap-based surveillance to immediately detect the effects of control measures. These findings underscore the importance of continuing pathogen testing yet re-evaluating spatial designs of trapping networks and control zones to better capture and respond to dynamic transmission.

Overall, this surveillance project tracked seasonal and spatial trends in mosquito populations, highlighting *Cx. pipiens* s.l. as the key vector, fitting the dogma of WNV transmission. Local *Cx. pipiens* s.l. picked up the virus through avian host feeding early in Maryland’s mosquito season, as seen through the peak vector index in July of both years. Transmission to humans coincided, with the first reported human cases occurring shortly afterward in August (health.maryland.gov 2024). Our results also convey that IDX enables efficient and timely identification and robust data curation as actionable insights for targeted mosquito control. Its use in Anne Arundel County allowed MDA to optimize abatement strategies, reduce WNV risks, and most importantly, promote public health outcomes. Ultimately, this work highlights the promise of smart surveillance platforms like IDX to improve vector control precision, bolster outbreak preparedness, and protect community health through faster, data-based decision-making.

## Data Availability

All data produced in the present work are contained in the manuscript

## Acknowledgements

We would like to acknowledge the Maryland Department of Health Laboratories Administration, Division of Molecular Biology - Molecular Diagnostics/Bioterrorism, including David Crum and colleagues, and the Anne Arundel County Department of Health, including Don Curtian and colleagues for their involvement in pathogen testing and reporting. This material is based upon work supported by the National Science Foundation under Grant No. 2322335, as well as funding from the Maryland Department of Agriculture. We would also like to acknowledge S. Jeanne Zastrow and Scott Larzelere for their contributions of trap site coordination and mosquito control efforts.

