## Supplemental Figures and Tables for "AI-Driven Surveillance of West Nile Virus in Maryland: Integrating Vector Identification with Environmental and Epidemiological Insights"

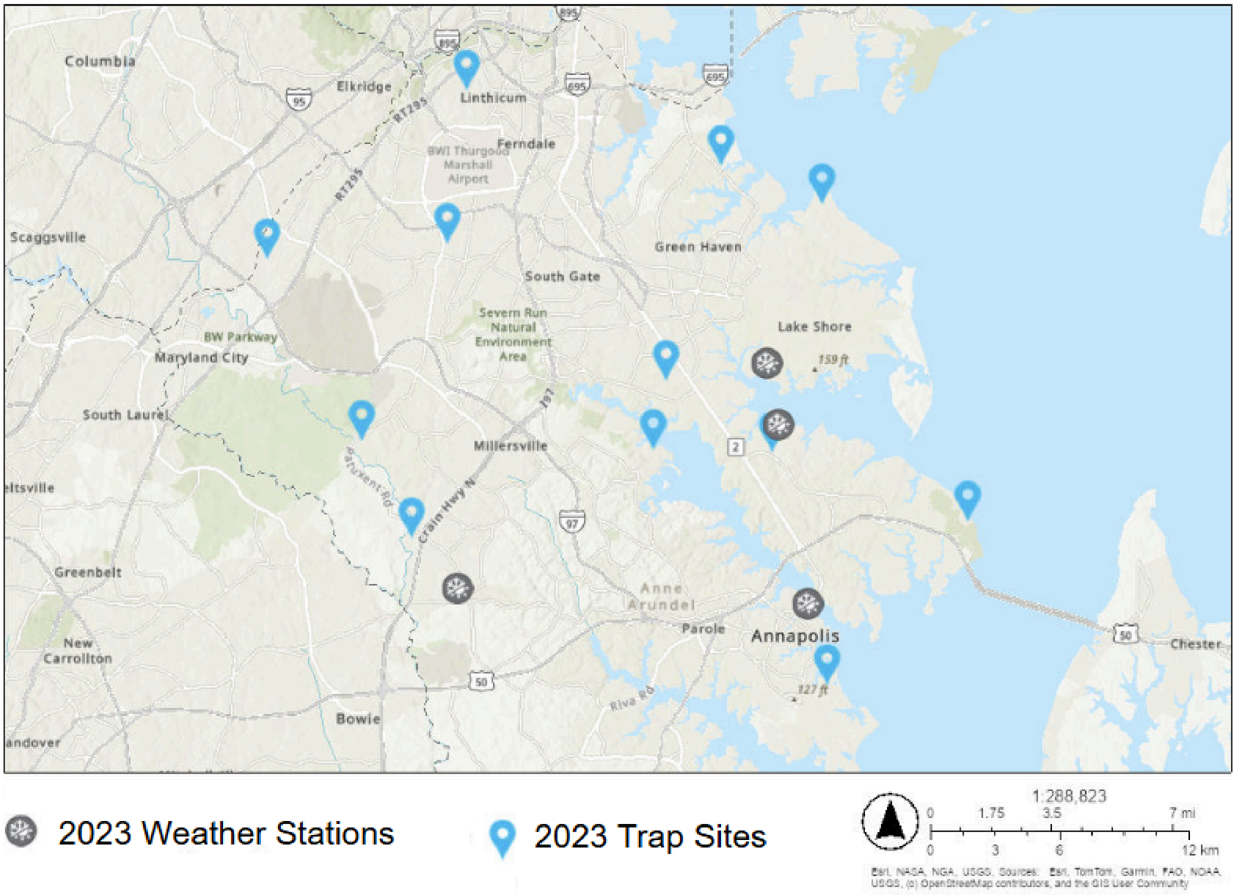


**Supplementary Figure S1.** 2023 MDA mosquito surveillance map. Twelve mosquito collection sites (depicted by blue location tags) and four weather stations (depicted by gray circles with sun and snowflakes icons) for 2023 arboviral surveillance in Anne Arundel County.

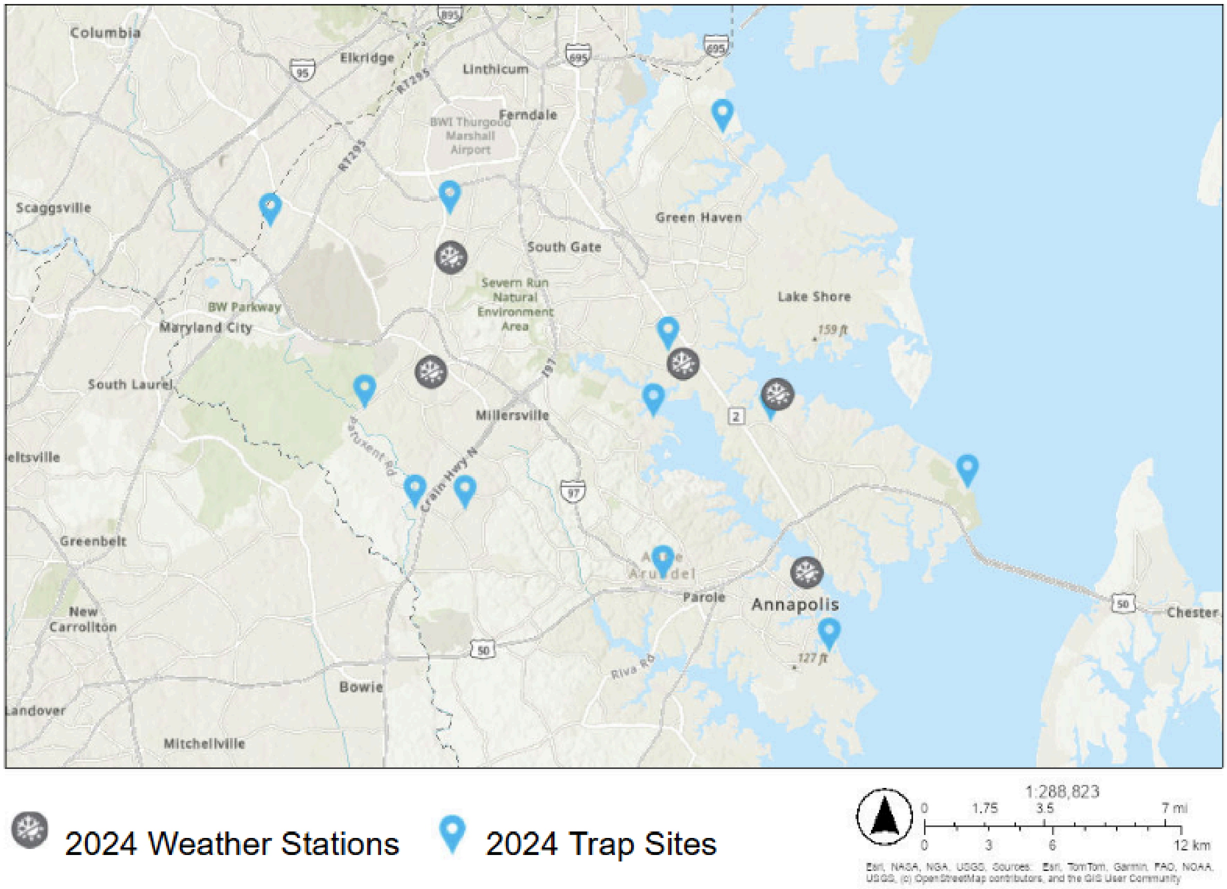


**Supplementary Figure S2.** 2024 MDA mosquito surveillance map. Twelve mosquito collection sites (depicted by blue location tags) and five weather stations (depicted by gray circles with sun and snowflakes icons) for 2024 arboviral surveillance in Anne Arundel County.

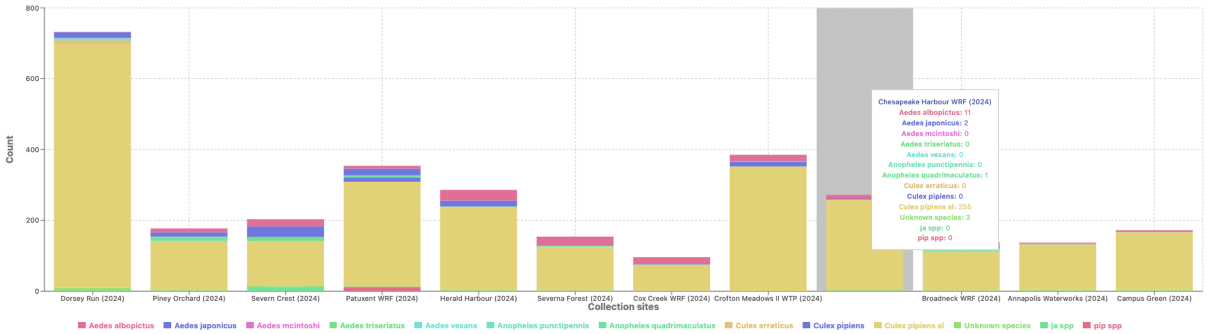


**Supplementary Figure S3**. Example of IDX Species Summary Among Collection Sites report. Imaged mosquito species graphed by the 12 collection sites during the 2023 surveillance period for MDA as seen in the IDX dashboard.

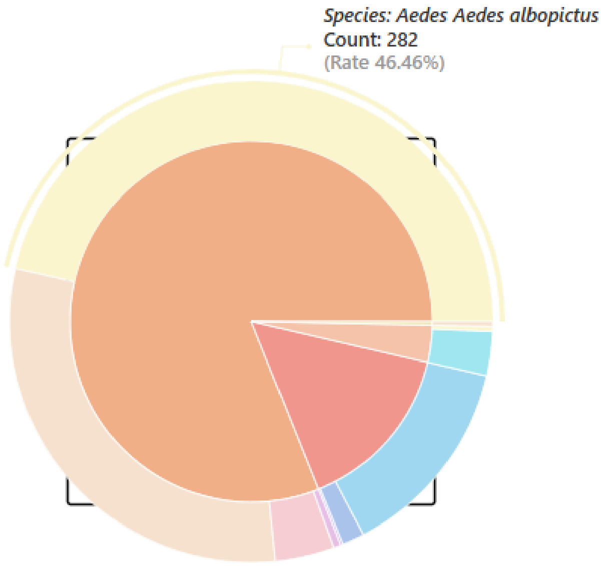


Legend

| Species | Count |
|---|---|
| *Aedes aedes albopictus* | 282 |
| *Aedes aedes japonicus* | 183 |
| *Anopheles anopheles quadrimaculatus* | 83 |
| *Aedes aedes triseriatus* | 24 |
| *Culex culex erraticus* | 18 |
| *Anopheles anopheles punctipennis* | 9 |
| *Aedes aedes vexans* | 3 |
| *Culex culex salinarius* | 2 |
| more... | 3 |
| Total | 607 |

**Supplementary Figure S4**. Example of IDX Species Summary report. Imaged mosquito species reported on a pie chart (left) with associated legend (right) as seen in the IDX dashboard.

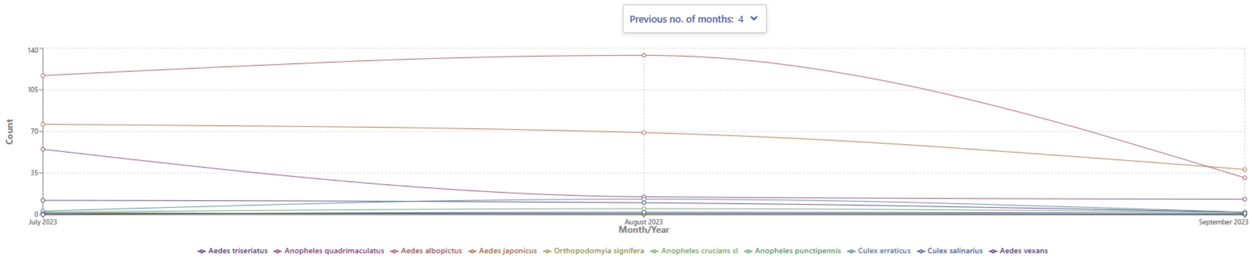


**Supplementary Figure S5.** Example of IDX Species Abundance by Month report. Imaged mosquito species graphed by month as seen in the IDX dashboard.

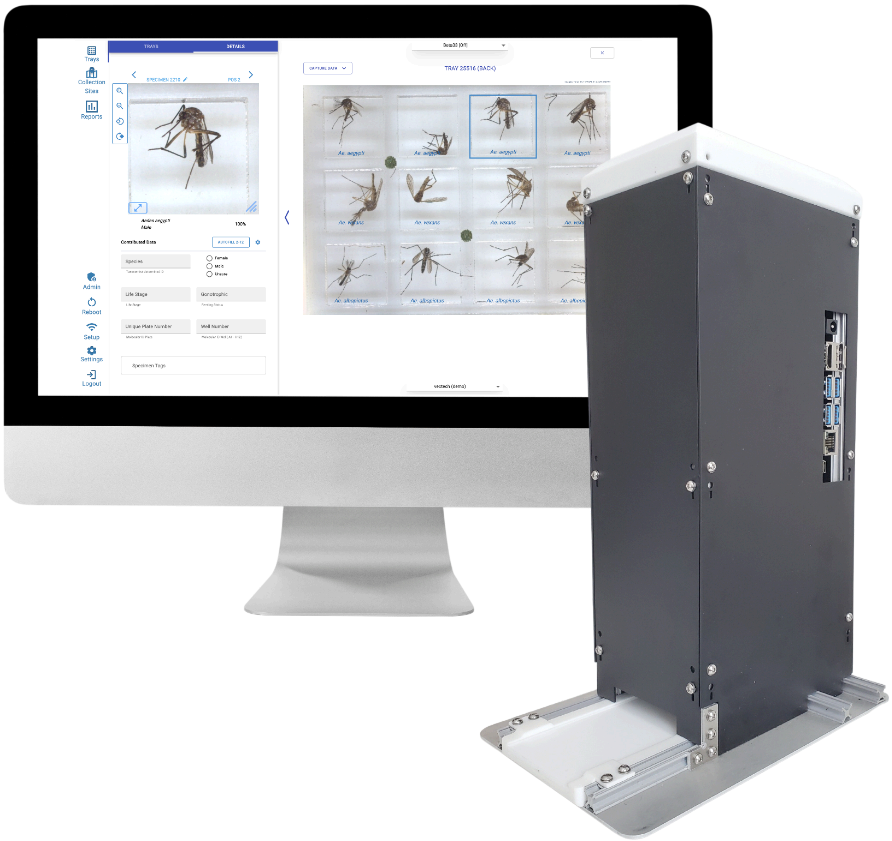


**Supplementary Figure S6**. Vectech IDX imaging device and dashboard. IDX (right) with online dashboard depicted on a computer screen (left) showing specimen images with algorithm species identification.

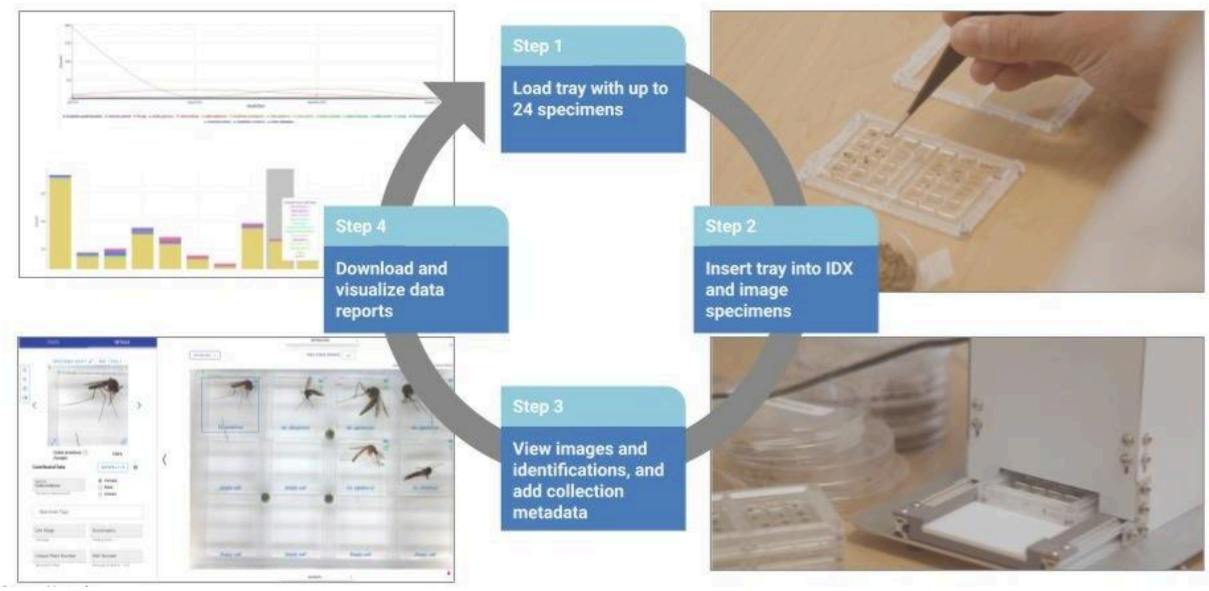


**Supplementary Figure S7**. IDX workflow diagram. Step 1 (top right) depicts the loading of individual mosquito specimens into IDX tray wells. Step 2 (bottom right) shows a tray loaded with specimens inserted in the IDX device. Step 3 (bottom left) details the IDX web-based dashboard as it assigns species and sex identification to imaged specimens within seconds. Step 4 (top left) includes detailed reports on data such as species abundance by collection site, collection method, date, etcetera.

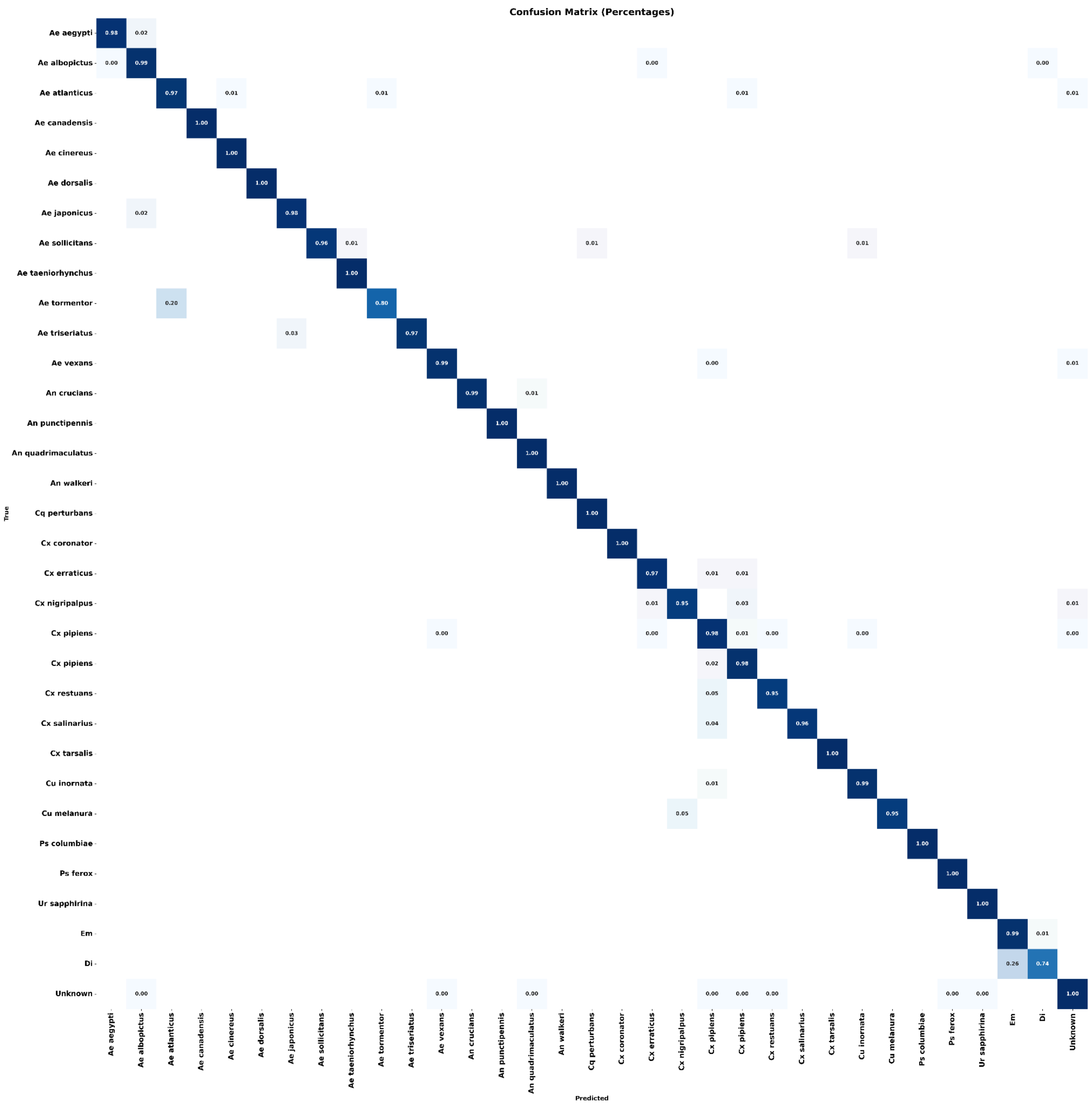


**Supplementary Figure S8.** Mid-atlantic confusion matrix on the latest IDX algorithm version consisting of 58 mosquito species. A confusion matrix is a visualisation table that shows classification error distributed among the species. The rows and columns depict the actual species name and predicted species name by the AI model, respectively. The diagonal shaded boxes show the accuracy with which a certain species is predicted correctly on a gradient (lighter blue for lower accuracy, darker blue for greater accuracy).

**Supplementary Table 1.** Pearson correlation coefficient (r) and corresponding p-values for relationship between William's mean mosquito abundance and both cumulative precipitation and average temperature at different weekly lag intervals (0-5) for 2023 and 2024.

| Week Lag | Year | Abundance vs. precipitation | Abundance vs. temperature |
|---|---|---|---|
| 0 | 2023 | r = -0.340, p = 0.429 | r = 0.577, p = 0.389 |
| | 2024 | r = -0.255, p = 0.378 | r = 0.074, p = 0.801 |
| 1 | 2023 | r = -0.302, p = 0.315 | r = 0.474, p = 0.102 |
| | 2024 | r = -0.265, p = 0.360 | r = -0.178, p = 0.541 |
| 2 | 2023 | r = 0.033, p = 0.914 | r = 0.131, p = 0.670 |
| | 2024 | r = -0.122, p = 0.678 | r = 0.043, p = 0.884 |
| 3 | 2023 | r = 0.564, p = 0.045 | r = -0.342, p = 0.253 |
| | 2024 | r = 0.302, p = 0.294 | r = -0.152, p = 0.604 |
| 4 | 2023 | r = 0.139, p = 0.651 | r = -0.562, p = 0.045 |
| | 2024 | r = 0.029, p = 0.921 | r = 0.114, p = 0.697 |
| 5 | 2023 | r = -0.121, p = 0.708 | r = -0.701, p = 0.009 |
| | 2024 | r = 0.349, p = 0.242 | r = -0.297, p = 0.325 |
